# Religious leaders’ perspectives on childhood immunization in Bauchi State, Nigeria: A Qualitative Study

**DOI:** 10.64898/2026.02.19.26346395

**Authors:** Auwal Abubakar, Stefano M Bertozzi, Adamu Mohammed, Rabiu Alhaji Suleh, Bakunawa Garba Bello, Suraj Musa Inuwa, Purnima Madhivanan, Maiya G Block Ngaybe, Olumayowa Adebayo, Ndola Prata, Doug Oman

## Abstract

**Introduction:** Childhood immunization is highly cost-effective, yet uptake is shaped by sociocultural and religious influences. In Bauchi State, Nigeria, coverage remains low (Penta3 58.2%; fully vaccinated 22.5% among children aged 12–23 months). Religious leaders shape community norms, but their perspectives on immunization in Bauchi are not well characterized. Guided by Oman and Syme’s religion/spirituality and health model, we explored religious leaders’ knowledge, beliefs, attitudes, practices and recommendations regarding childhood immunization.

**Methods:** Between December 2022 and January 2023, we conducted semi-structured interviews with 22 religious leaders (all men; 21 Muslim, 1 Christian) purposively sampled across Bauchi State’s Local Government Areas. Interviews were conducted in Hausa or English, audio recorded, translated where necessary and transcribed. Sampling continued until thematic saturation. Data were analyzed using codebook thematic analysis informed by Braun and Clarke’s analytic phases and organized according to the pre-specified domains of knowledge, beliefs, attitudes, practices and recommendations. Ethical approval was obtained, and all participants provided verbal informed consent.

**Results:** Most leaders described immunization as preventive and compatible with religious teachings. Many reported that observed child health benefits reinforced support and that earlier skepticism had shifted after religious and scientific explanation and lived experience. Persistent misinformation was reported, especially fertility-related and population control narratives. Leaders described three recurring influence practices: visible role modelling, sermon-based messaging aligned with scripture, and community mobilization through religious gatherings and support during outreach activities. Respectful health worker engagement and reliable service delivery appeared to strengthen trust and community uptake.

**Conclusions:** Religious leaders in Bauchi State may be strategic partners for improving vaccine acceptance. Programs should consider structured engagement with religious leaders, bidirectional rumor tracking and response, and support for frontline health workers. Future research should include more diverse leaders and link engagement to measurable outcomes.

**KEY MESSAGES:** *What is already known on this topic:* Childhood immunization coverage remains suboptimal in northern Nigeria, and religious leaders can influence vaccine-related beliefs and uptake, yet their perspectives and practical roles are not well characterized in Bauchi State.

*What this study adds:* Among 22 interviewed religious leaders in Bauchi State, most viewed childhood immunization as compatible with religious teachings and described using role modelling, sermon-based messaging, and community mobilization to support uptake, while also reporting persistent fertility-related and population control rumors and service delivery barriers.

*How this study might affect research, practice or policy:* Immunization programs may benefit from structured engagement with religious leaders, locally credible rumor response systems, and parallel improvements in respectful and reliable service delivery.

## INTRODUCTION

Childhood immunization is a cornerstone of global public health and among the most cost-effective health interventions.^1^ Immunization averts an estimated 3.5–5.0 million deaths annually.^1^ In Nigeria, routine childhood vaccination coverage remains suboptimal. In the 2024 Nigeria Demographic and Health Survey (NDHS), national coverage of three doses of pentavalent vaccine (penta3) was 53.4% and coverage in Bauchi State was 58.2% among children aged 12–23 months.^2^ However, only 19.8% of children nationally and 22.5% in Bauchi were fully vaccinated according to the national schedule.^2^ These levels remain below the IA2030 target of ≥90% coverage for the third dose of a DTP-containing vaccine (reported as DTP3; in Nigeria, measured as Penta3).^3^ Given these gaps, understanding sociocultural drivers of uptake is essential. Although vaccination is a biomedical intervention, acceptance and uptake are shaped by sociocultural context, including religious beliefs and practices.^4^ Specifically, religious beliefs and authorities can influence perceptions, ethical evaluations, including concerns about vaccine ingredients, and community norms around vaccination.^4^

Religious leaders can influence vaccination uptake in either direction. In northern Nigeria, religious and political leaders’ skepticism contributed to the 2003–2004 polio vaccine boycott, during which parents were publicly urged to refuse vaccination amid contamination and fertility-related allegations, disrupting immunization activities and exacerbating mistrust.^5^ Conversely, subsequent polio eradication efforts in northern Nigeria engaged Muslim clerics to counter misconceptions and promote uptake.^6^ A survey of Muslim scholars in Pakistan similarly found generally positive attitudes toward polio immunization and described engagement of religious scholars to address polio-related myths.^7^ These contrasts underscore the dual potential of religious leadership to shape public health outcomes.

Given their influence, the Bauchi State Primary Health Care Development Agency (BSPHCDA), with support from partners, developed a routine immunization community partnership strategy in 2016 that leveraged traditional and religious leaders.^8^ Despite this effort, completion of the full national schedule remains low (22.5% in Bauchi in the NDHS 2024),^2^ underscoring the importance of understanding where religious leaders’ perspectives fall along a supportive to obstructive spectrum to inform future engagement strategies. However, there is limited qualitative evidence focused specifically on religious leaders’ own perspectives in Bauchi State, including how they understand childhood immunization, reconcile it with religious teachings, and translate endorsement into concrete mobilization practices. This information is needed to optimize faith-based engagement strategies.

Accordingly, guided by the leadership pathway in our conceptual framework (Figure 1), we qualitatively assessed religious leaders’ knowledge, beliefs, attitudes, practices and recommendations (KBAPR) regarding childhood immunization in Bauchi State.

**Figure 1.**
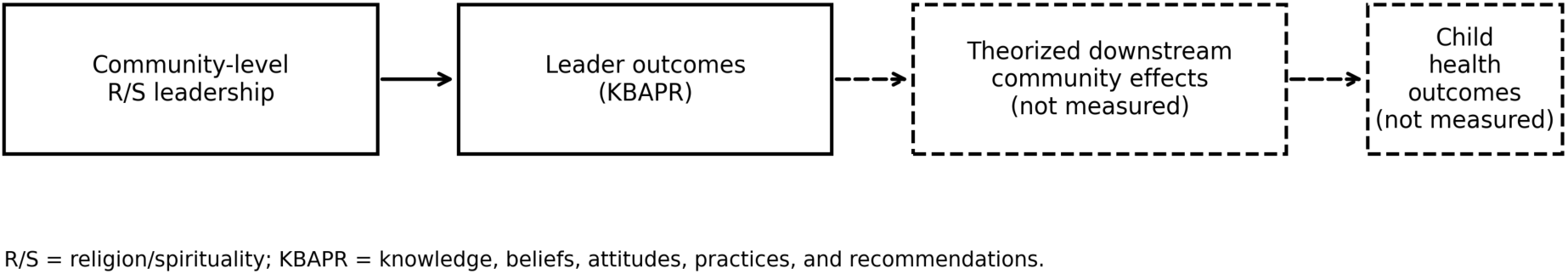
Conceptual framework adapted from Oman and Syme (2018). This figure isolates the community-level leadership pathway and indicates theorized downstream effects that were not measured

### THEORY AND CONCEPTUAL FRAMEWORK

Guided by a leadership pathway adapted from Oman and Syme (2018),^9^ we pre-specified leadership as the focal community-level religion/spirituality (R/S) dimension. In this model, community-level R/S is multidimensional, including leadership, beliefs, practices, feelings, and coping, and we examined the leadership dimension. We pre-specified leader-level outcomes: knowledge, beliefs, attitudes, practices, and recommendations (KBAPR), as the primary outcomes and focused our analysis on this upstream leadership component of the pathway. We did not evaluate downstream community vaccination behaviors or hypothesized biological mechanisms in this study.

## METHODS

### Study design

We used a semi-structured interview guide to explore religious leaders’ perspectives on childhood immunization in Bauchi State, Nigeria. The guide captured demographics (age, ethnicity, religion, employment, education, and income) and elicited leaders’ knowledge, beliefs, attitudes, practices, and recommendations (KBAPR). In the final guide, probes covered sources of information; perceived benefits and challenges; community reception; leaders’ roles in campaigns; relationships with health workers; strategy preferences (house-to-house vs routine immunization) and rationales; and recommendations for communities, providers, and peers. The guide was KBAPR-informed and aligned with our conceptual focus on community-level religious leadership. Reporting follows the COREQ 32-item checklist (Supplementary File 1).^10^ The guide was pilot-tested with two non-study religious leaders (one Imam, one Christian cleric). Based on pilot feedback, we simplified phrasing, reordered demographic items, and added probes on sources of information and relationships with health workers. We also added probes to capture and clarify changes over time in leaders’ positions. When a leader described a shift in views (toward or away from support), we asked follow-up questions about factors influencing the change and whether peers’ views had also changed. Pilot interviews were excluded from analysis.

### Sample description

The study population comprised religious leaders serving congregations in Bauchi State, Nigeria. For this study, a religious leader was defined as an adult (≥18 years) currently recognized by a mosque or church congregation, or its community, as its leader. Eligible participants resided in, or primarily served within, Bauchi State and could complete an interview in Hausa or English. Individuals not holding a current leadership role, those unable to participate because of language or cognitive barriers, or those who declined or withdrew consent were not included. All interviewees were men, reflecting the predominance of men among formal religious leaders in the study sampling frame. The final sample (N = 22) included 21 Muslim leaders and 1 Christian leader, consistent with the composition of the eligible sampling frame. Participant characteristics are summarized in Table 1.

**Table 1.** Sociodemographic characteristics of participants (religious leaders) (N = 22)

| Characteristic | Value, n (%) or mean (SD) |
| --- | --- |
| **Age, years** |  |
| Mean (SD) | 51.6 (10.1) |
| Range | 36–73 |
| **Ethnicity, n (%)** |  |
| Hausa | 10 (45.5) |
| Fulani | 6 (27.3) |
| Kanuri | 2 (9.1) |
| Sayawa | 1 (4.5) |
| Bolewa | 1 (4.5) |
| Bajari | 1 (4.5) |
| Baduguri | 1 (4.5) |
| **Religion, n (%)** |  |
| Islam | 21 (95.5) |
| Christianity | 1 (4.5) |
| **Employment, n (%)** |  |
| Civil servant | 13 (59.1) |
| Farmer | 6 (27.3) |
| Retired | 3 (13.6) |
| **Education (highest attained), n (%)** |  |
| Qur'anic education only | 5 (22.7) |
| National Certificate in Education (NCE) | 6 (27.3) |
| Diploma (OND/HND) | 5 (22.7) |
| Bachelor's degree | 4 (18.2) |
| NCE plus Diploma | 2 (9.1) |
| **Annual income (NGN), median (IQR)<sup>a</sup>** | NGN 636,000 (NGN 561,000–NGN 951,000)<br>≈USD 1,413 (USD 1,247–USD 2,113)<sup>b</sup> |
| **Formal involvement in immunization activities, n (%)** |  |
| Yes | 8 (36.4) |
| No | 14 (63.6) |

**Notes:** Values are n (%) unless otherwise indicated. Percentages are calculated with denominator N = 22 and may not sum to 100% due to rounding; Education categories reflect reported educational attainment; NCE plus Diploma (OND/HND) indicates participants reporting both credentials; OND/HND = Ordinary/Higher National Diploma; NGN = Nigerian naira; USD = United States dollars; SD = standard deviation; IQR = interquartile range; <sup>a</sup>Income reported by 20/22 participants; <sup>b</sup> USD conversions use NGN450 per USD (January 2023, at data collection);

### Sampling, recruitment and sample size

We used geographically stratified purposive sampling from a Bauchi State Primary Health Care Development Agency (BSPHCDA) roster of 415 mosque and church leaders. To ensure statewide coverage, we targeted at least one leader from each of Bauchi State’s 20 local government areas (LGAs), and the final sample included at least one leader from each LGA. Ten alternates were preselected to mitigate nonresponse.

Potential participants were contacted by telephone by the principal investigator (PI), screened against eligibility criteria, and invited to a face-to-face interview at the Immunization Plus and Malaria Progress by Accelerating Coverage and Transforming Services (IMPACT) project office. Telephone interviews were offered when travel constraints precluded in-person participation. However, all enrolled participants completed face-to-face interviews. Participants received travel-distance–based reimbursement of NGN10,000–NGN15,000 (approximately US$22–34) at the time of data collection.

Sample size was determined by thematic saturation. We tracked the emergence of new codes and subthemes after each interview using a running saturation table (interviews by emerging codes). After interview 22, no new codes emerged, and the codebook was finalized. Recruitment was therefore closed at N = 22, with no refusals and no dropouts.

### Data collection

All interviews were conducted in person by the PI at the IMPACT project office in Bauchi LGA from December 2022 to January 2023. The location was chosen for accessibility and privacy. To reduce social desirability bias, the interviewer emphasized that there were no right or wrong answers and reiterated confidentiality. Verbal informed consent, including consent to audio-record, was obtained prior to each interview. Interviews were conducted in Hausa or English based on participant preference, were audio-recorded, and only the interviewer and participant were present. Interviews lasted an average of 40 minutes (range, 22–78 minutes). At the end of each interview, we conducted brief member-checking by summarizing key points to participants and inviting corrections or clarifications. Hausa interviews were translated and transcribed into English by the PI, a native Hausa speaker. A research team member whose mother tongue is Hausa and who is fluent in English independently verified the translations against the audio, and discrepancies were resolved by consensus. No repeat interviews were conducted, and transcripts were not returned to participants. Quotations presented in the results section were lightly edited for readability; ellipses indicate omissions and bracketed text indicates clarifications.

### Researcher reflexivity

Interviews were conducted by the PI (male, MBBS, MPH, MSc.), who was a doctoral student in public health at the University of California, Berkeley at the time of data collection and is currently affiliated with the University of Arizona. He trained in qualitative interviewing and thematic analysis through graduate coursework and supervised practice. The PI is a native Hausa speaker fluent in English and has prior professional experience in immunization programs in northern Nigeria, a background that may have oriented him toward a generally favorable view of immunization.

To mitigate potential bias, the PI used a KBAPR-informed, semi-structured guide, emphasized neutrality and the absence of right or wrong answers, and stated independence from service- delivery programs. The PI also worked with a second analyst who independently coded transcripts. Discrepancies were resolved by consensus. There were no preexisting personal relationships with participants. Sampling used a BSPHCDA roster with initial telephone contact. At recruitment and consent, participants were told the PI’s university affiliation, study aims, and that participation was voluntary and confidential.

### Analysis

We used a codebook thematic analysis approach, informed by Braun and Clarke’s analytic phases,^11^ and managed data in ATLAS.ti, version 22.0.1. Two analysts (the PI and a co-investigator) independently coded transcripts in two cycles. In cycle 1, we performed open, inductive coding. In cycle 2, we organized codes into a hierarchical codebook anchored in our a priori KBAPR domains: knowledge, beliefs, attitudes, practices, and recommendations, while retaining inductive subthemes.

Intercoder reliability was assessed on a random 25% subset of transcripts (n = 6 of 22) using Cohen’s kappa across the full code set (κ = 0.73; percent agreement = 86%). Domain-specific κ ranged from 0.62 to 0.84, indicating moderate to substantial agreement. Discrepancies were discussed and resolved by consensus, and the codebook was refined iteratively throughout analysis. We iteratively reviewed candidate themes against coded extracts and the full dataset, then defined and named final themes. Themes are presented with representative quotations (participant IDs only).

To enhance trustworthiness, we strengthened credibility through pilot testing and iterative refinement of the interview guide, in-interview member checking (summarizing key points and inviting corrections and clarifications), and analyst triangulation (two independent coders with consensus discussions). We supported dependability and confirmability by using a documented codebook, assessing intercoder agreement, and tracking thematic saturation using a running saturation table. Transferability was supported by geographically stratified purposive sampling across all 20 LGAs and detailed reporting of participant and setting characteristics.

### Patient and public involvement

Neither patients nor the public were involved in the design, conduct, reporting, or dissemination plans of this study.

### Data security and retention

Audio files and transcripts were stored on a password-protected, encrypted computer with access restricted to the study team. Audio recordings were deleted after transcription was verified for accuracy. De-identified transcripts are retained in accordance with the approved protocol for potential secondary analyses.

## RESULTS

### Sociodemographic characteristics

We interviewed 22 religious leaders (all men). Their mean age was 51.6 years (SD 10.1; range 36–73). Hausa and Fulani were the most represented ethnicities. Most participants identified as Muslim. Civil servant was the most common occupation, followed by farmer. Educational attainment ranged from Qur’anic education only (no formal Western schooling) to tertiary qualifications. Among the 20 participants reporting income, the median (IQR) annual income was NGN 636,000 (NGN 561,000–NGN 951,000), approximately USD 1,413 (USD 1,247– USD 2,113). Full sociodemographic distributions are summarized in Table 1.

### Thematic Analysis

Findings were grouped into five pre-specified domains: Knowledge, Beliefs, Attitudes, Practices, and Recommendations (KBAPR). Table 2 summarizes subthemes, key insights, and participant frequency labels (few/some/many/most). Representative quotations are provided in Supplementary File 2.

**Table 2.**
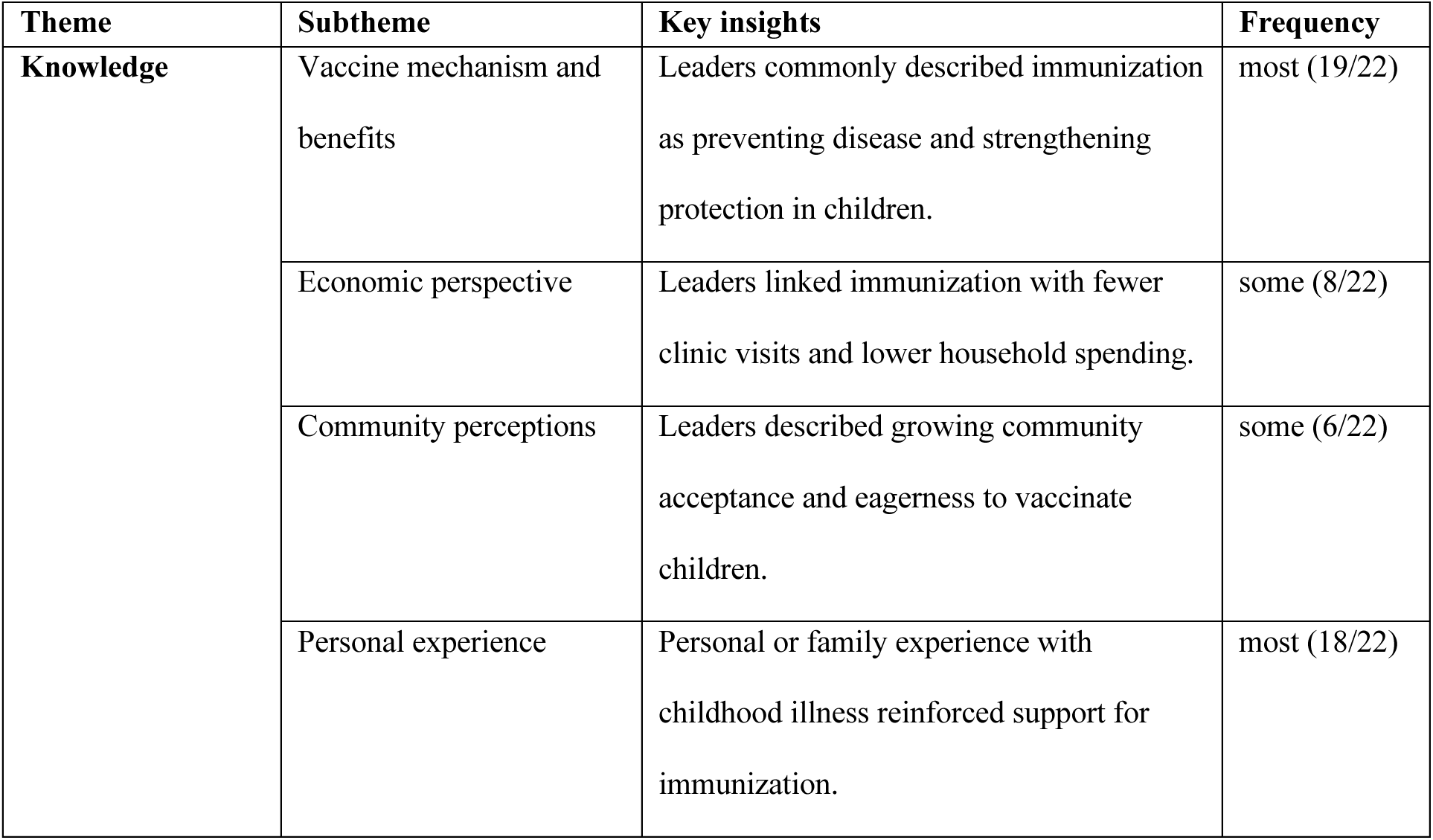
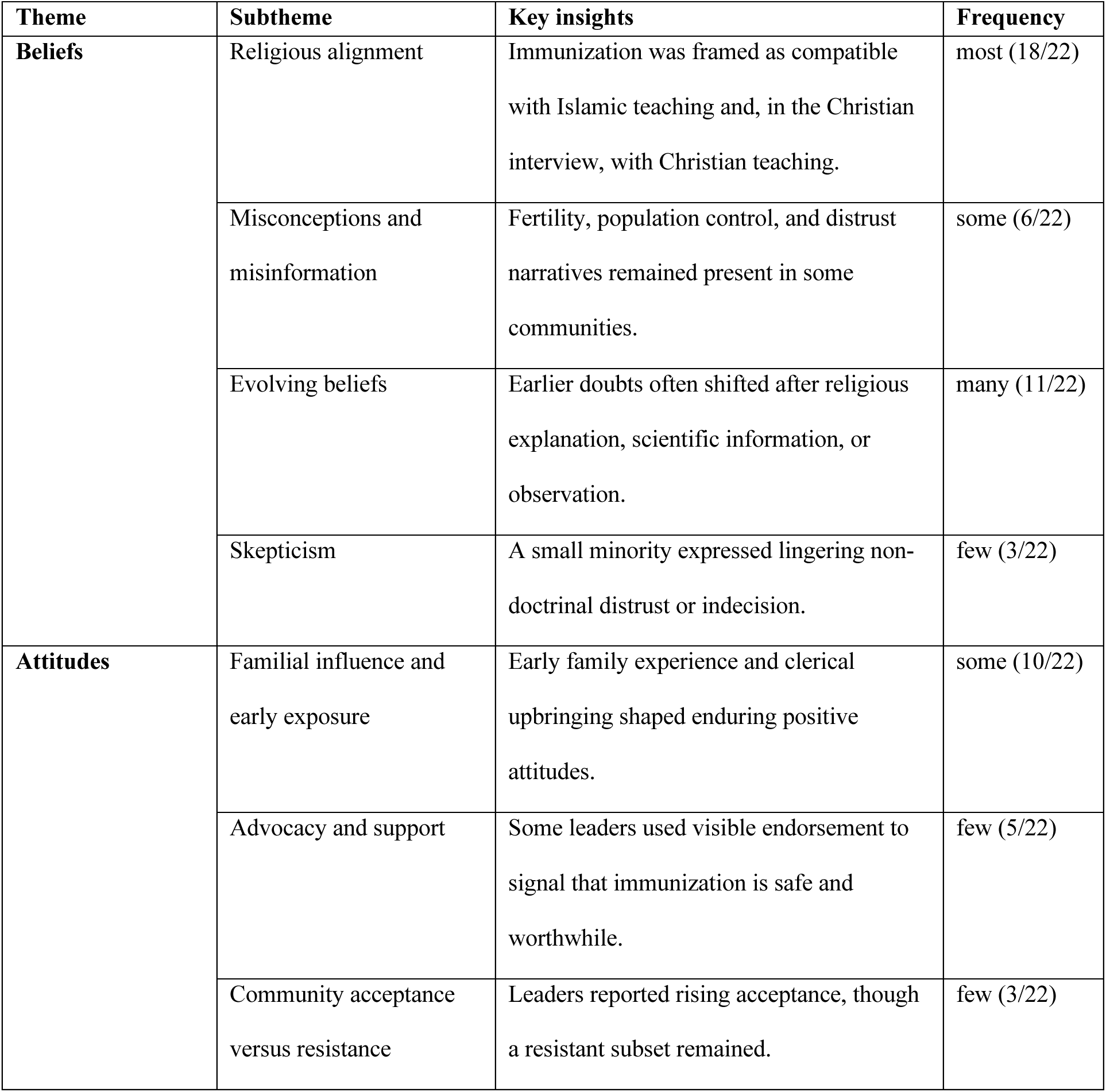
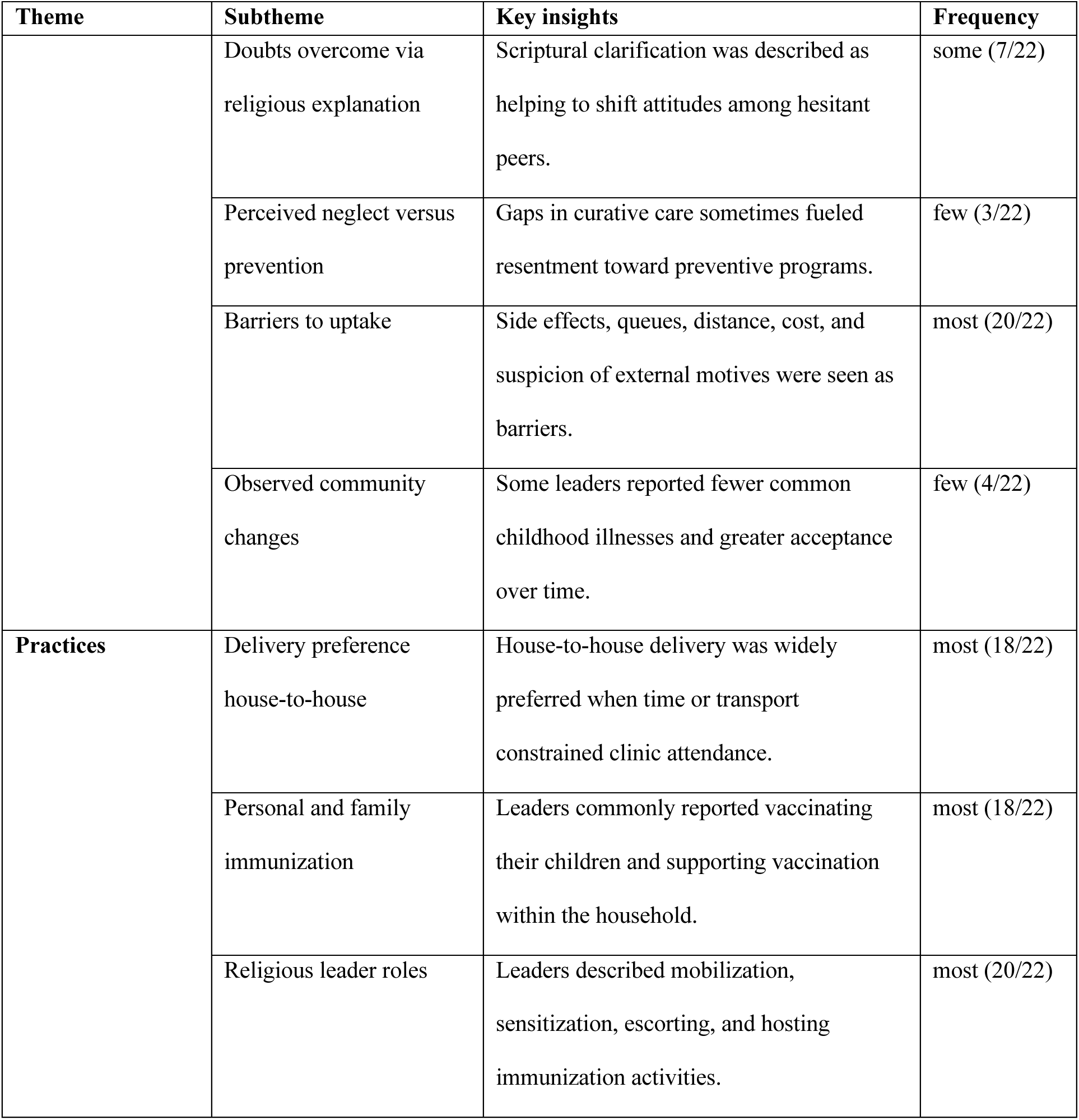
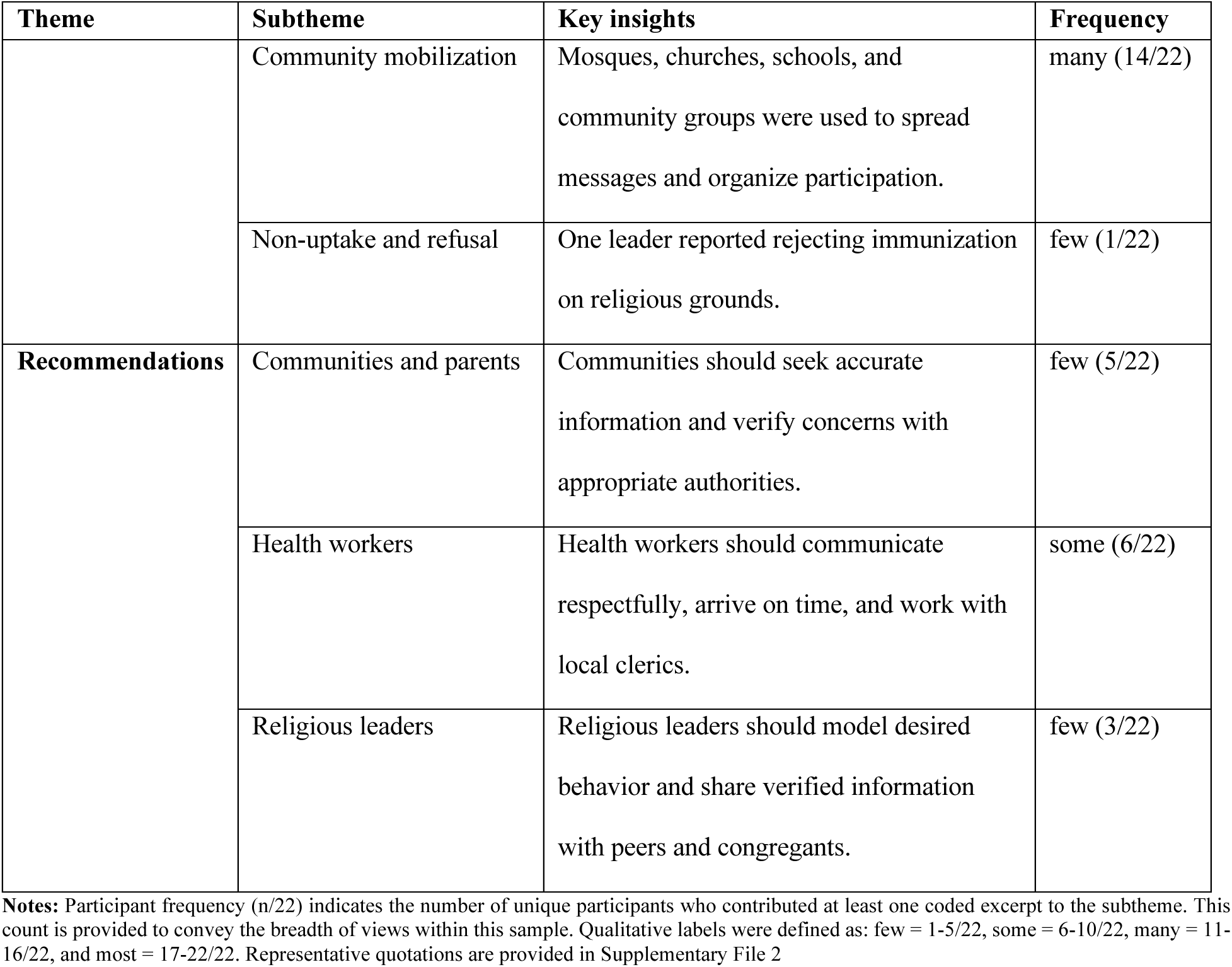
Summary of themes, subthemes, key insights, and participant frequency (N = 22)

### Knowledge about childhood immunization

*As summarized in Table 2,* religious leaders described varying levels of knowledge about childhood immunization, with accounts clustering around understanding of mechanism, perceived health and economic benefits, and community perceptions. Most recognized vaccines as preventive measures that protect children from disease. One leader articulated this understanding as a remedy administered before illness manifests:

> *“What I understand is that it is a remedy for a perceived disease that is approaching. So, it is given to protect them from being affected… they help in lowering the effects of disease from childhood.” (Participant 10).*

Some leaders emphasized perceived economic benefits to families when children remain healthy.

Leaders often contrasted the perceived financial strain of curative care with the relative stability they associated with vaccinated households. As one leader observed:

> *“Unlike those who were not vaccinated, their parent must struggle and visit hospital and spend a lot of money, while the one who was vaccinated… recovers within short time.” (Participant 8).*

When describing community perceptions, some leaders pointed to visible signs of increasing acceptance. For example, one leader described a crowded maternity clinic and interpreted this as evidence of high uptake:

> *“I visited the maternity… I saw the crowd of women that were waiting to immunize their children… to me it implies the acceptance is high.” (Participant 4).*

Overall, leaders perceived fewer hospital visits and declines in some childhood illnesses and often attributed these changes to immunization based on their observations.

### Beliefs on childhood immunization

Belief statements reflected participants’ interpretations of religious doctrine and practice (Table 2). Most leaders described immunization as compatible with disease prevention and with religious teachings. The single Christian religious leader interviewed expressed a similar view. Several linked immunizations to scriptural and prophetic encouragement to seek remedies and framed acceptance as consistent with faith-based responsibility. One Muslim leader cited a Hadith encouraging treatment and analogized an early-life prophetic practice involving date extract as protective:

> *“The Prophet Muhammad SAW was saying ‘tadawa’u ya ibadallah…’ (seek treatment, O servants of Allah) … And The Prophet SAW said if a child is delivered, before he or she swallows anything, an extract of date should be dropped in his or her mouth… that drops of date extract is vaccination…” (Participant 8).*

A Christian leader similarly emphasized that immunization does not contradict Christian teaching

> *“Christianity does not disagree with childhood immunization… I have never come across any part of my scripture which teaches against such a thing.” (Participant 4)*

One leader conceptualized immunization through the Islamic principle of protection of life

> *“There is no disagreement between Islamic religion and immunization, it is known as hifzul nafsi which is protection of lives. One must protect the lives of his child, and to also prevent him against diseases…” (Participant 7)*

Alongside supportive beliefs, leaders also described persistent misinformation, including early rumors linking immunization to fertility reduction:

> *“Initially we were not supporting, because it was said it’s for the reduction of reproduction.” (Participant 9).*

Overall, leaders described strong religious endorsements of health seeking alongside ongoing misinformation and distrust in some communities.

### Attitudes toward childhood immunization

Most religious leaders expressed supportive attitudes, commonly grounded in upbringing and perceived compatibility with faith (Table 2). Many previously held reservations, often linked to misinformation, were reported to have shifted after religious discussions and dialogue. One respondent illustrated this shift by citing a specific community example:

> *“There is someone I know. He is a religious leader, but he doesn’t vaccinate his children… When I ask him why he changed and now accepting, he told me [it] was as a result of influence by some religious leaders.” (Participant 3).*

Some skepticism was framed around delivery approach, especially house-to-house (H2H) vaccination, which was sometimes interpreted as suspicious precisely because it targets healthy children:

> *“People are saying that it is dangerous… they wonder why free medicine is being brought H2H, even to healthy people… there might be a hidden agenda to it.” (Participant 19).*

Leaders also linked hesitancy to perceived inconsistencies in health system responsiveness, for example comparing limited access to drugs with the availability of free vaccines:

> *“Others complain that there are no drugs in the hospital and drugs are costly … but despite the cost these vaccines are given for free.” (Participant 10).*

Overall, leaders described largely supportive attitudes that often strengthened over time through dialogue and religious framing, while residual hesitancy was commonly linked to mistrust, concerns about house-to-house delivery, and perceived inconsistencies in broader health system responsiveness.

### Practices related to childhood immunization

Beyond statements of support, leaders described how they translated endorsement into routine, practical engagement. For example, one leader described using Friday sermons and other gatherings to announce upcoming activities and persuade caregivers to participate:

> *“If I come to the Friday prayer and give a sermon, I will convince people that they should go and receive this injection today… if I come to the mosque or our Islamic school, I don’t have to wait for Friday. I will announce this.” (Participant 11).*

Another leader described relaying campaign information through church structures, briefing ward-level leaders and mandating announcements so members would be aware and participate:

> *“We call those who are under our leadership … we brief them and then sometimes we mandate them to announce it in churches so that people will be aware… and participate.” (Participant 4).*

Overall, leaders actively translated their support into practice by leveraging their pulpits, organizational hierarchies, and routine religious gatherings to mobilize community participation.

### Recommendations for childhood immunization

Leaders directed recommendations to three audiences.

Communities and parents: Seek accurate information from governmental and health sources; share verified information with peers; and view immunization as preventive and compatible with faith:

> *“Immunization is something that is accepted by Allah and his prophet peace be upon him and is in line with the teaching of the prophet, it is there in Islam, the prophet did hijama, which is cupping, since the prophet himself did it, we as Muslims emulate him.” (Participant 7)*

Health workers and programs: Use courteous, patient communication; be punctual; and coordinate visits in advance with rural religious leaders:

> *“…use kind words and be courteous.” (Participant 17)*

Religious leaders (peers): Peer-to-peer learning to address misconceptions, and dissemination of accurate knowledge from authoritative sources.

> *“As leaders, we make ourselves available in order to show good examples for the people and to be a role model for followership. This is because there is the need to practice what you preach… as a leader, the eyes of many people are on you to hear your own opinion, and it’s your opinion that makes people either accept or reject whatever is presented to them.” (Participant 1);*
>
> *“Every religious leader who researches and understands that something is not prohibited, should come out and share the knowledge by creating awareness for other leaders and his people.” (Participant 3)*

Across audiences, recommendations converged on accurate information, respectful engagement, coordination with faith actors, and leadership by example.

## DISCUSSION

In this qualitative study of 22 religious leaders in Bauchi State, Nigeria, leaders generally endorsed childhood immunization and grounded their support in religious teachings and reported observations of perceived child health benefits. Supportive views may partly reflect prior exposure to immunization programs in the state, including structured engagement with religious leaders since 2016.^8^ In our sample, 8 of 22 leaders reported formal involvement in immunization activities.

Leaders also described persistent misinformation, most notably fertility-related and population control rumors, and they emphasized that community acceptance was strengthened when immunization services were delivered respectfully and reliably. Across interviews, leaders described three recurring influence practices: (i) Visible role modelling, (ii) Sermon-based messaging aligned with scripture, and (iii) Community mobilization through announcements, sensitization, and coordination with outreach activities.

These findings align with evidence from Nigeria and other settings that faith leaders can amplify vaccine acceptance, while distrust and misinformation may also sustain hesitancy.^5,6,12–14^ In Indonesia, a qualitative study of religious and community leaders similarly found that leaders viewed childhood vaccination as compatible with Islamic principles and positioned themselves as bridges between immunization programs and their communities, though they emphasized the need for adequate information and early, genuine engagement.^14^ We extend prior work by specifying, from leaders’ accounts, conditions under which clerical influence may be amplified or attenuated: the salience of rumors, service accessibility and reliability, and the quality of health worker engagement. Qualitative research in Bauchi and Cross River States has documented that poor health worker attitudes and deficient communication skills undermine caregiver trust, and that religious and traditional leaders were identified as useful and acceptable channels of vaccination communication, particularly during campaigns.^15^ A similar pattern was reported in Sokoto State, where poor interpersonal communication between health workers and caregivers was a major demand-side barrier and communities themselves recommended engaging trusted religious leaders to bridge the trust deficit.^16^ We also distinguish pulpit messaging from interpersonal counselling and community mediation. Leaders suggested that visible endorsement (for example, accompanying families for routine immunization or publicly encouraging uptake during services) may help normalize vaccination and reduce perceived risk, whereas long queues, limited staffing and negative service experiences may inadvertently reinforce skepticism, highlighting the interdependence between community persuasion and program reliability.

Leaders demonstrated a generally strong understanding of immunization benefits and frequently framed acceptance as consistent with religious duty and child welfare. However, misconceptions remained salient in community narratives and were described as re-emerging through social networks and public messaging. A qualitative study engaging religious leaders across Ghana in polio vaccine promotion similarly found that leaders and community members held culturally and religiously influenced explanations for disease and emphasized that mobilization strategies developed in genuine collaboration with religious leaders, rather than using them as passive conduits, were more likely to gain traction.^17^ Several leaders reported shifts over time from earlier skepticism to support after religious clarification, additional information and observed outcomes.

These accounts suggest that engagement strategies may be most effective when they combine credible religious framing with timely, comprehensible responses to circulating rumors.

Mapped to Oman and Syme’s model of R/S and health, our results primarily reflect leader-level outcomes (knowledge, beliefs, attitudes, practices and recommendations) within the community-level leadership pathway. We did not assess downstream congregant intentions, vaccination behaviors or coverage, and therefore cannot infer effects attributable to religious leadership. Leaders’ accounts nonetheless highlight that service-delivery frictions, such as travel burden, long waits, and limited staff responsiveness, may blunt the potential effect of religious endorsement, and several leaders described attempting to mitigate these barriers through coordination with health personnel and advising during community gatherings.

This study has several strengths, including a piloted semi-structured guide, reporting informed by COREQ, geographically stratified purposive sampling to support statewide breadth, and an analytic process involving independent coding with consensus resolution.

Limitations include potential selection bias due to reliance on a health-agency roster, possible social desirability bias because interviews were conducted in an office linked to an immunization project, and limited diversity in gender and denomination (21 Muslim and 1 Christian leader; all men), which constrains transferability to female religious leaders and Christian or other non- Muslim faith contexts. Because data were collected only from leaders, we could not assess congregant perspectives or verify behavioral effects of leaders’ actions.

Programmatically, findings support pragmatic strategies that leverage leaders’ existing influence while strengthening service experience. Potential actions include co-developing sermon and message kits (scriptural anchors plus concise myth-busting points), supporting visible role modelling through planned participation in immunization events and strengthening bidirectional rumor tracking between leaders and primary health care teams to enable timely, locally credible responses. A community-based misinformation management program in Niger State, Nigeria, which trained fellows from diverse backgrounds, including religious organizations, in rumor identification, community listening, and response, illustrates one possible model for operationalizing such strategies in related settings.^18^ These community-facing strategies may be strengthened when paired with respectful, punctual and reliable service delivery, including attention to waiting time and health worker–client interaction.

Future research should purposively recruit dissenting or less engaged leaders and include more diverse religious leadership (including women and non-Muslim leaders), link leader engagement to congregant outcomes using mixed methods, and evaluate integrated leader–health worker engagement packages using pragmatic designs and implementation outcomes.

## CONCLUSION

In this qualitative study in Bauchi State, most religious leaders endorsed childhood immunization and described using visible role modelling, sermon-based messaging, community mobilization and engagement during outreach activities to promote acceptance, while acknowledging persistent fertility-related and conspiracy rumors. Leaders’ perceived influence appeared contingent on the local rumor environment and the reliability and respectfulness of service delivery. Because this leaders-only qualitative study did not measure congregant outcomes, we cannot infer effects on vaccination behavior or coverage. Programs may benefit from structured engagement with religious leaders, including co-developed sermon and message kits and bidirectional rumor tracking. Such efforts should be implemented alongside strategies to strengthen the service experience and support frontline health workers, and evaluated using measurable implementation and uptake indicators.

## Supporting information

COREQ (COnsolidated criteria for REporting Qualitative research) Checklist

## STATEMENTS

### Ethics statements

#### Consent for publication

Participants consented to publication of de-identified quotations.

#### Ethics

The study received ethical approval from the Bauchi State Health Research Ethics Committee (Bauchi State Ministry of Health; Protocol No. BSMOHREC/069/2022; NHREC Protocol No. NHREC/03/11/198/2021/69; expedited review, minimal risk). All participants provided verbal informed consent, including consent for audio recording, before the interview. Participant reimbursements were set in advance based on travel distance and were not contingent on responses.

### Author contributorship

Auwal Abubakar (AA) conceived the study, developed the interview guide, led study implementation, conducted all interviews, managed and curated the data, led the coding and thematic analysis, and drafted the manuscript.

Doug Oman (DO) made major contributions to study conception and design and substantively revised the manuscript.

Stefano M Bertozzi (SMB) and Ndola Prata (NP) provided academic supervision and methodological guidance and critically reviewed and revised the manuscript for important intellectual content.

Olumayowa Adebayo (OA) served as the second coder for intercoder reliability and contributed to refinement of the codebook and interpretation.

Adamu Mohammed (AM), Rabiu Alhaji Suleh (RAS), Bakunawa Garba Bello (BGB), Suraj Musa Inuwa (SMI), Purnima Madhivanan (PM) and Maiya G Block Ngaybe (MGBN) provided contextual expertise and critical review and provided feedback on the manuscript. All authors reviewed and approved the final version. AA is the guarantor.

### AI use statement

The lead author used ChatGPT (OpenAI) during manuscript preparation to suggest edits for grammar, phrasing, and clarity. All AI-assisted suggestions were critically reviewed, edited, and verified by the authors, who take responsibility for the final content.

### Competing interests

The authors declare no competing interests.

### Funding

This research was supported by the University of California, Berkeley, Center for African Studies, Andrew and Mary Thompson Rocca Fellowship and the Fogarty International Center of the National Institutes of Health under Award Number D43TW010540, through the Global Health Emerging Scholars program. The funders had no role in study design, data collection and analysis, decision to publish, or manuscript preparation. The content is solely the responsibility of the authors and does not necessarily reflect the official views of the National Institutes of Health.

### Data availability statement

Data are available upon reasonable request. Deidentified interview transcripts, the interview guide, and study materials are available from the corresponding author, subject to ethics approval, institutional requirements, and a data use agreement. Requests should be directed to the corresponding author at.

## ACKNOWLEDGMENTS

We thank the Berkeley School of Public Health for their support throughout this research. We also thank the Bauchi State Primary Health Care Development Agency for providing the list of religious leaders and the IMPACT office for private locations for conducting interviews.

This research would not have been possible without the participation of the religious leaders in Bauchi State, who generously shared their time, insights, and experiences. Their invaluable contributions have greatly enriched this work.

## Supplementary File 2

**Table 2.**
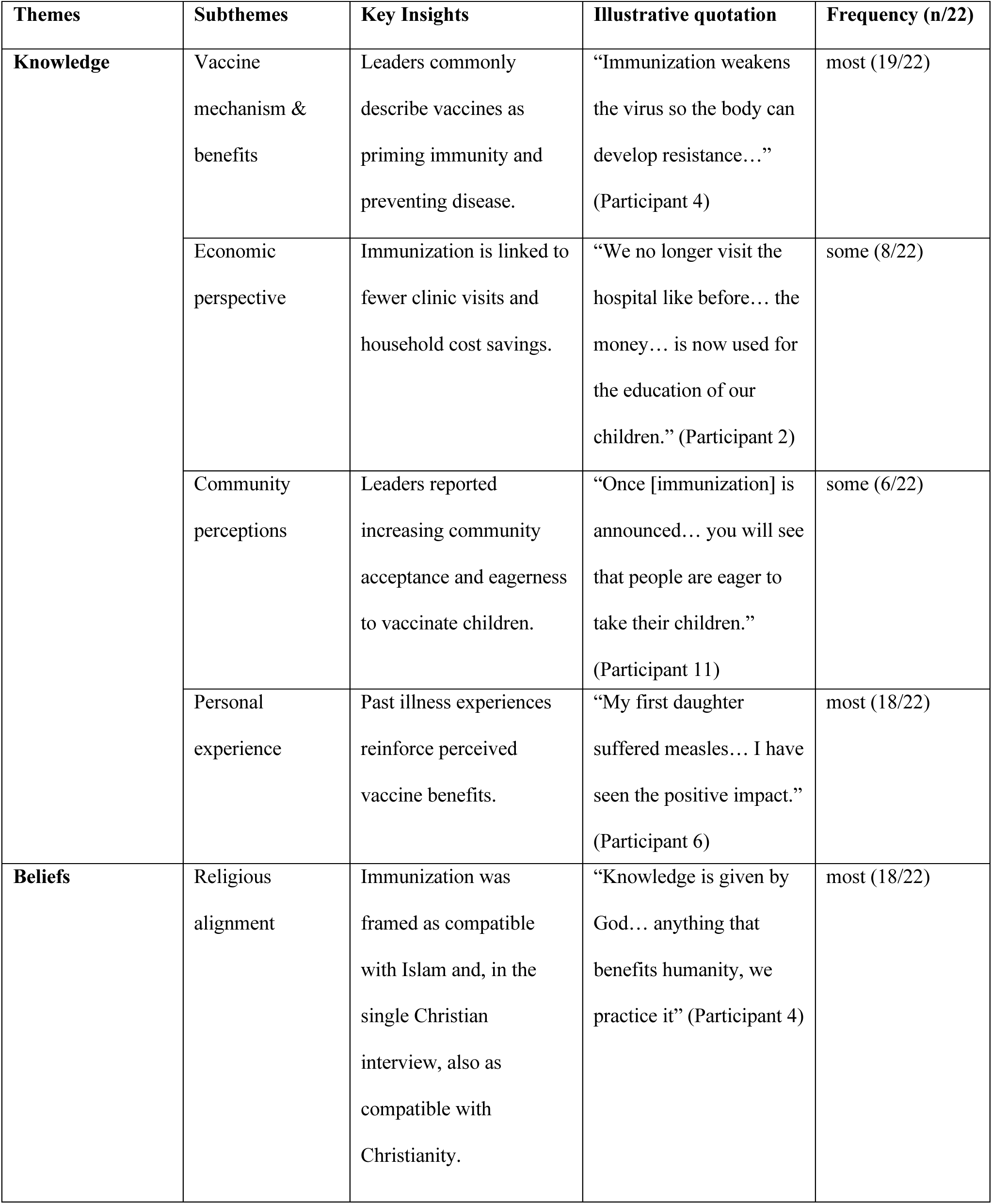
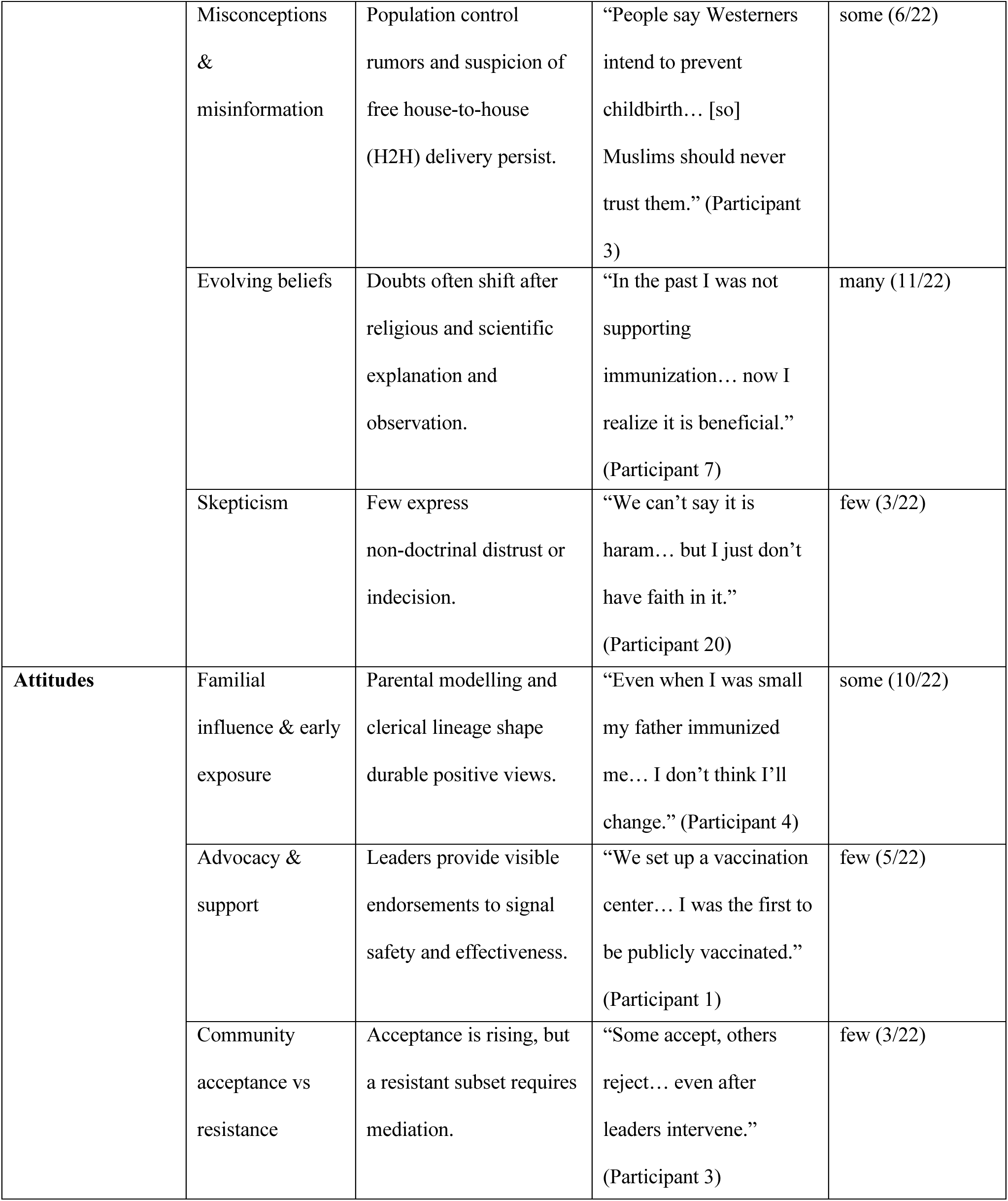
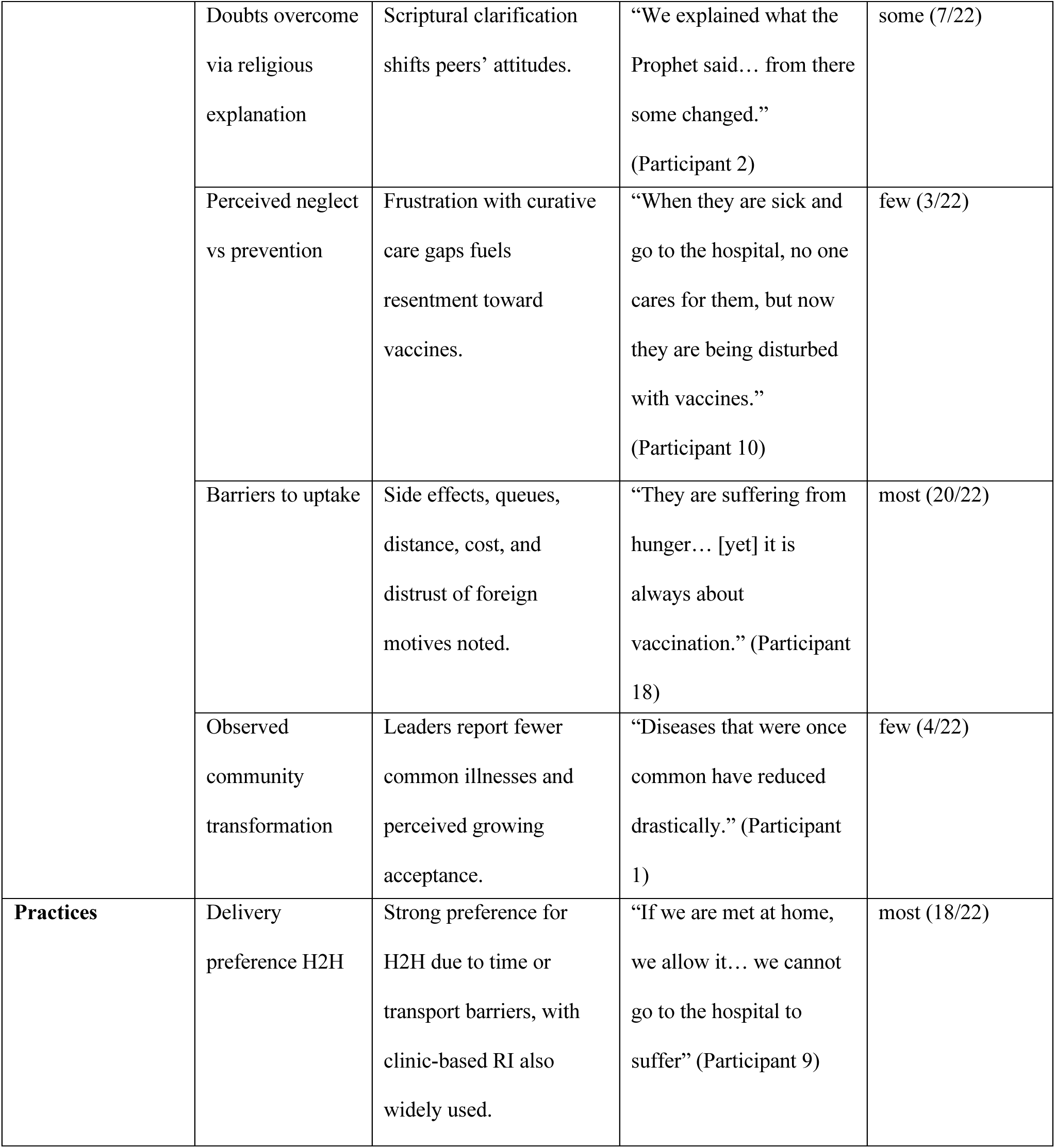
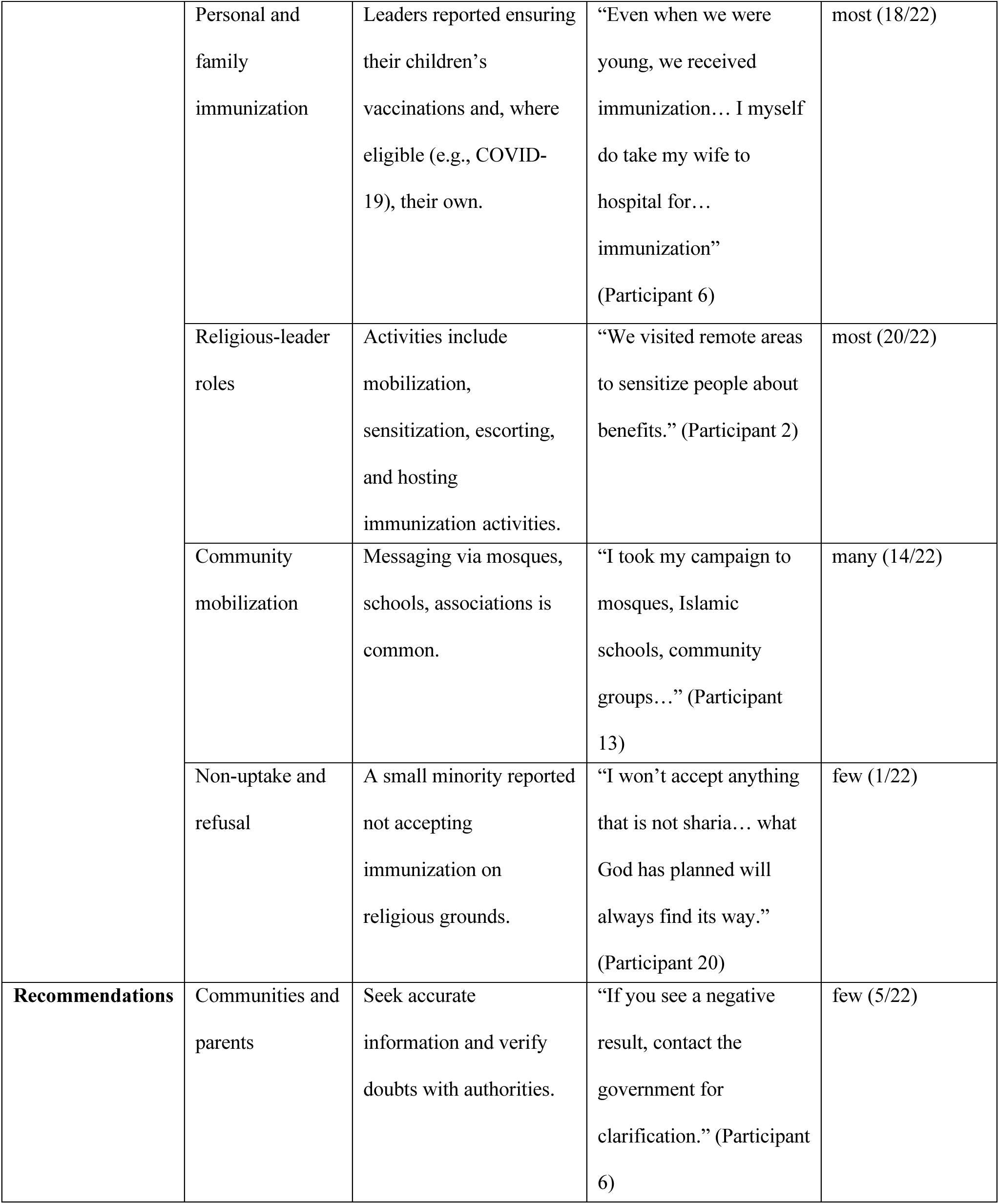
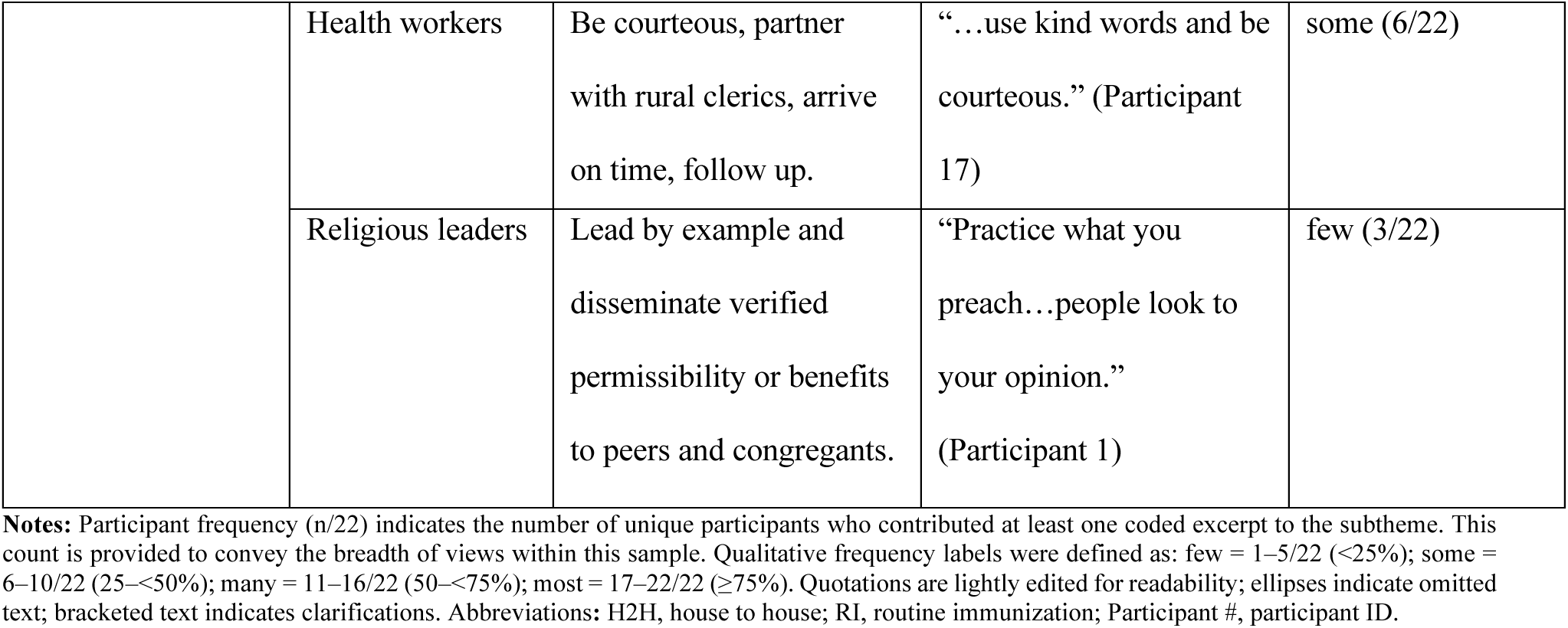
Thematic Analysis overview of Themes, Subthemes, Key Insights, and Representative Quotes (N = 22)

| Themes | Subthemes | Key Insights | Illustrative quotation | Frequency (n/22) |
| --- | --- | --- | --- | --- |
| **Knowledge** | Vaccine mechanism & benefits | Leaders commonly describe vaccines as priming immunity and preventing disease. | “Immunization weakens the virus so the body can develop resistance...”<br>(Participant 4) | most (19/22) |
|  | Economic perspective | Immunization is linked to fewer clinic visits and household cost savings. | “We no longer visit the hospital like before... the money... is now used for the education of our children.” (Participant 2) | some (8/22) |
|  | Community perceptions | Leaders reported increasing community acceptance and eagerness to vaccinate children. | “Once [immunization] is announced... you will see that people are eager to take their children.”<br>(Participant 11) | some (6/22) |
|  | Personal experience | Past illness experiences reinforce perceived vaccine benefits. | “My first daughter suffered measles... I have seen the positive impact.”<br>(Participant 6) | most (18/22) |
| **Beliefs** | Religious alignment | Immunization was framed as compatible with Islam and, in the single Christian interview, also as compatible with Christianity. | “Knowledge is given by God... anything that benefits humanity, we practice it” (Participant 4) | most (18/22) |
|  | Misconceptions & misinformation | Population control rumors and suspicion of free house-to-house (H2H) delivery persist. | “People say Westerners intend to prevent childbirth... [so] Muslims should never trust them.” (Participant 3) | some (6/22) |
|  | Evolving beliefs | Doubts often shift after religious and scientific explanation and observation. | “In the past I was not supporting immunization... now I realize it is beneficial.” (Participant 7) | many (11/22) |
|  | Skepticism | Few express non-doctrinal distrust or indecision. | “We can’t say it is haram... but I just don’t have faith in it.” (Participant 20) | few (3/22) |
| **Attitudes** | Familial influence & early exposure | Parental modelling and clerical lineage shape durable positive views. | “Even when I was small my father immunized me... I don’t think I’ll change.” (Participant 4) | some (10/22) |
|  | Advocacy & support | Leaders provide visible endorsements to signal safety and effectiveness. | “We set up a vaccination center... I was the first to be publicly vaccinated.” (Participant 1) | few (5/22) |
|  | Community acceptance vs resistance | Acceptance is rising, but a resistant subset requires mediation. | “Some accept, others reject... even after leaders intervene.” (Participant 3) | few (3/22) |
|  | Doubts overcome via religious explanation | Scriptural clarification shifts peers' attitudes. | "We explained what the Prophet said... from there some changed."<br>(Participant 2) | some (7/22) |
|  | Perceived neglect vs prevention | Frustration with curative care gaps fuels resentment toward vaccines. | "When they are sick and go to the hospital, no one cares for them, but now they are being disturbed with vaccines."<br>(Participant 10) | few (3/22) |
|  | Barriers to uptake | Side effects, queues, distance, cost, and distrust of foreign motives noted. | "They are suffering from hunger... [yet] it is always about vaccination." (Participant 18) | most (20/22) |
|  | Observed community transformation | Leaders report fewer common illnesses and perceived growing acceptance. | "Diseases that were once common have reduced drastically." (Participant 1) | few (4/22) |
| **Practices** | Delivery preference H2H | Strong preference for H2H due to time or transport barriers, with clinic-based RI also widely used. | "If we are met at home, we allow it... we cannot go to the hospital to suffer" (Participant 9) | most (18/22) |
|  | Personal and family immunization | Leaders reported ensuring their children's vaccinations and, where eligible (e.g., COVID-19), their own. | "Even when we were young, we received immunization... I myself do take my wife to hospital for... immunization"<br>(Participant 6) | most (18/22) |
|  | Religious-leader roles | Activities include mobilization, sensitization, escorting, and hosting immunization activities. | "We visited remote areas to sensitize people about benefits." (Participant 2) | most (20/22) |
|  | Community mobilization | Messaging via mosques, schools, associations is common. | "I took my campaign to mosques, Islamic schools, community groups..." (Participant 13) | many (14/22) |
|  | Non-uptake and refusal | A small minority reported not accepting immunization on religious grounds. | "I won't accept anything that is not sharia... what God has planned will always find its way."<br>(Participant 20) | few (1/22) |
| **Recommendations** | Communities and parents | Seek accurate information and verify doubts with authorities. | "If you see a negative result, contact the government for clarification." (Participant 6) | few (5/22) |
|  | Health workers | Be courteous, partner with rural clerics, arrive on time, follow up. | “...use kind words and be courteous.” (Participant 17) | some (6/22) |
|  | Religious leaders | Lead by example and disseminate verified permissibility or benefits to peers and congregants. | “Practice what you preach...people look to your opinion.” (Participant 1) | few (3/22) |

**Notes:** Participant frequency (n/22) indicates the number of unique participants who contributed at least one coded excerpt to the subtheme. This count is provided to convey the breadth of views within this sample. Qualitative frequency labels were defined as: few = 1–5/22 (<25%); some = 6–10/22 (25–<50%); many = 11–16/22 (50–<75%); most = 17–22/22 (≥75%). Quotations are lightly edited for readability; ellipses indicate omitted text; bracketed text indicates clarifications. Abbreviations: H2H, house to house; RI, routine immunization; Participant #, participant ID.

## Notes

### Competing Interest Statement

The authors have declared no competing interest.

### Author Declarations

The study received ethical approval from the Bauchi State Health Research Ethics Committee (Bauchi State Ministry of Health; Protocol No. BSMOHREC/069/2022; NHREC Protocol No. NHREC/03/11/198/2021/69; expedited review, minimal risk)

### Summary of Updates

This revised version updates the supplementary materials. Table 2 has been moved from the main manuscript to a supplementary file, and the COREQ checklist has been added as a supplementary file.

